# Genetic architecture and polygenic prediction of self-reported response to selective serotonin reuptake inhibitors in individuals with depression: a genome-wide meta-analysis

**DOI:** 10.64898/2026.11.11.26367161

**Authors:** Michelle Kamp, Madhurbain Singh, Ke Hu, Chris Wai Hang Lo, Cameron J. Watson, Brittany L. Mitchell, Jodi T. Thomas, Gerome Breen, Jonathan R.I. Coleman, Sang-Hyuck Lee, Oliver Pain, 23andMe Research Institute, AMBER Research Team, AGDS Research Team, GLAD Study Consortium, the NIHR BioResource, Cathryn M. Lewis

## Abstract

Selective serotonin reuptake inhibitors (SSRIs) are first-line treatments for depression, but response is highly variable, and no robust genetic predictors have been identified. We conducted a genome-wide association meta-analysis of self-reported SSRI response in 62,879 individuals of European ancestry with depression (72.6% responders) across six cohorts (seven datasets), followed by gene-set, transcriptomic, proteomic, drug-target and polygenic score (PGS) analyses. SNP-based heritability was modest but significant (h² = 3.1%, SE = 1.4%) with no genome-wide significant variants detected. Gene-set analyses implicated gamma-aminobutyric acid (GABA)ergic and glutamatergic neurotransmission, and a gene involved in glutamate transport (*SLC17A6*) showed consistent sub-threshold evidence across pathway, transcriptomic and proteomic analyses. Drug-target enrichment for hypnotics, anxiolytics and general anesthetics further supported the GABAergic signal. A leave-one-out PGS from the discovery meta-analysis was associated with self-reported SSRI response in independent cohorts (meta-analysis OR=1.05, 95% CI 1.02–1.08, p=0.002). Higher PGS for depression, attention-deficit/hyperactivity disorder (ADHD), anxiety, post-traumatic stress disorder (PTSD), autism, addiction and schizophrenia were associated with lower odds of reported SSRI response, with ADHD, PTSD, autism and addiction remaining after adjustment for depression PGS. In contrast, educational attainment PGS was associated with better SSRI response. Self-reported SSRI response captures genetic insights relevant to antidepressant response, implicating GABAergic and glutamatergic neurotransmission. Individuals with higher genetic loading for psychiatric conditions report poorer response, and higher genetic loading for educational attainment PGS a better reported response. Although larger studies are needed to identify specific variants, self-report offers a practical, patient-centered and scalable route to collect the larger samples required.

## 1. Introduction

Major depressive disorder (MDD) is among the most prevalent and disabling conditions globally (1). Selective serotonin reuptake inhibitors (SSRIs) are a first-line pharmacological treatment (2) and the most widely prescribed antidepressant class (3), yet response is highly variable. Clinician-rated remission over 6-12-week trials occurs in around one-third of individuals (4), whereas up to 75% self-report benefit (5). This gap may reflect differences in outcome definition (remission versus perceived benefit), trial duration, recall bias, or how clinicians and patients judge meaningful improvement, and understanding it may provide insight into the biological mechanisms underlying SSRI response.

Genetic variation contributes to differences in antidepressant treatment outcomes (6–9), with single nucleotide polymorphism-based heritability estimated at 13% (7). As with many traits in psychiatry, antidepressant treatment response is highly polygenic (6–8,10), and genetic liability to other psychiatric disorders has been associated with poorer outcomes (11). To date, genome-wide association studies (GWAS) have found no robustly replicated loci. A recent five-cohort meta-analysis of SSRI response identified a single novel locus by combining electronic health record-based, self-reported and clinical trial response phenotypes, but combining phenotypes trades larger samples against greater heterogeneity (8).

Self-reported response mitigates this trade-off. A comparable self-report question across studies provides consistent ascertainment at larger sample sizes than clinical phenotypes allow. Collected via large-scale questionnaires, self-report is patient-centered and scalable, and may capture aspects of treatment outcome not routinely recorded in clinical settings (5). Despite recall bias, the resulting sample size gains may increase power for locus discovery and improve genetic architecture estimates.

One explanation for the disparity between clinically assessed and self-reported response is that self-report captures improvement over longer periods than trials, as real-world antidepressant use often extends over several years (12). If so, genetic and transcriptomic signals for self-reported response should be enriched in pathways implicated in antidepressant action and depression biology.

Here, we present the largest GWAS meta-analysis of self-reported SSRI response. We characterize its genetic architecture, test this hypothesis through pathway and drug-class enrichment analyses, and assess whether genetic liability to other psychiatric conditions is associated with reported outcome.

## 2. Methods

### 2.1 Primary GWAS

Primary GWAS of self-reported SSRI response were conducted in three cohorts: All of Us Research Program (AoU; (13)), Genetic Links to Anxiety and Depression (GLAD; (14)), and UK Biobank (UKB; (15)) (Supplementary Note1). Binary SSRI response was derived from a retrospective, self-reported effectiveness question answered for each SSRI taken. SSRIs included citalopram, fluoxetine, paroxetine and sertraline in all cohorts, plus escitalopram and fluvoxamine in GLAD. For participants rating multiple SSRIs, majority positive ratings were classified as response and majority negative ratings as non-response. Conflicting ratings (e.g., one positive and one negative) were excluded. Analyses were limited to participants with lifetime MDD, defined by self-reported diagnoses (GLAD) or DSM-5 criteria applied to Composite International Diagnostic Interview -Short Form (CIDI-SF; AoU and UKB) (16). Participants reporting bipolar disorder, schizophrenia, schizoaffective disorder or psychosis were excluded. Analyses were restricted to participants genetically similar to European-ancestry reference individuals (EUR-like) due to small sample sizes in other ancestry groups (Supplementary TableS2).

In each cohort, GWAS was conducted using REGENIE (17), following sample- and variant-level quality control (Supplementary Note1). Logistic regression tested associations between SSRI response and sequenced genotypes (AoU, UKB) or imputed dosages (GLAD) of common variants (MAF >0.01, INFO >0.7 in GLAD), adjusting for age at recruitment, genetic sex, the first six genetic principal components (PCs), and cohort-specific technical factors. REGENIE’s approximate Firth correction was applied at p-values <0.01.

### 2.2 Genome-wide meta-analyses

In addition to the primary GWAS (AoU, GLAD and UKB), summary statistics of self-reported SSRI response were obtained from published GWAS in the Australian Genetics of Depression Study [AGDS; (18)], Estonian Biobank [EstBB; (19)], and 23andMe Research Institute [23andMe; (20,21)], as previously described (8) (Supplementary Note2).

Quality-controlled summary statistics were harmonized and meta-analyzed using an inverse-variance-weighted fixed-effect (FE) model in bcftools v1.22 (22) with score plugins +munge, +liftover and +metal ((23); https://github.com/freeseek/score#run-meta-analysis), using the - -overlap option to correct for sample overlap. As a sensitivity analysis, Han and Eskin’s modified random-effects (RE2) meta-analysis was implemented using METASOFT ((24); https://hanlab-snu.github.io/METASOFT/). SNP-level heterogeneity between cohorts was assessed using I² and Cochran’s Q statistics for SNPs in ≥3 cohorts (Supplementary Note3, Note4). Genome-wide and suggestive significance were defined as p <5×10⁻⁸ and p <5×10⁻⁶, respectively.

### 2.3 Functional annotation and gene-based analyses

Independent genomic loci were defined in FUMA SNP2GENE (v1.8.2) (25) at the suggestive threshold (p <5×10⁻⁶), using the 1000 Genomes Phase 3 European reference panel (1KGP3 EUR), and SNPs within these loci were positionally mapped to protein-coding genes (Ensembl v102). MAGMA (v1.08) (26) was used for gene-based association analyses (20,068 protein-coding genes), competitive gene-set analysis (17,012 MSigDB gene sets and GO terms) and tissue-expression analysis (54 specific and 30 general GTEx v8 tissues), with Bonferroni correction within each analysis. Positionally mapped genes were characterized using GENE2FUNC. Analyses were repeated on RE2 summary statistics (Supplementary Note5).

### 2.4 SNP-based heritability and genetic correlations

Meta-analytic SNP-based heritability (*SNP-h²*) was estimated using LD score regression (LDSC; (27)) and converted to the liability scale (28), assuming population prevalence equals sample prevalence (0.726). Genetic correlations (*rg*) with 17 traits, spanning psychiatric conditions, personality, antidepressant response, BMI and educational attainment, were estimated using bivariate LDSC (Supplementary Note5, Supplementary TableS3). Benjamini-Hochberg FDR correction (q < 0.05) was applied across with significant SNP-h² and a successfully estimated *rg* (Supplementary Note5 and Table S3).

### 2.5 Polygenic scoring analyses

Polygenic scores (PGS) were calculated with SBayesRC in GenoPred (v2.3.3) (29), using GCTB LD matrices from EUR-like UKB participants and restricted to HapMap3 SNPs (30–32); Supplementary Note5). To evaluate the predictive performance of a self-reported SSRI-response PGS, we constructed leave-one-out (LOO) PGS, excluding each target cohort (AGDS, AoU, GLAD) from the meta-analysis in turn. Associations were tested using logistic regression in unrelated participants, adjusting for age, sex and the first six PCs (AoU, GLAD) or sex and the first ten PCs (AGDS). Variance explained was calculated as the incremental Nagelkerke’s R² over a covariates-only model and converted to the liability scale (33), assuming a population prevalence of 0.726. Cohort-specific odds ratios (per SD of the PGS) were combined using fixed-effect inverse-variance meta-analysis (*metafor* in R; (34)), with heterogeneity assessed using Cochran’s Q and I².

To examine shared genetic architecture, PGS for the traits in the genetic correlation analysis (Supplementary TableS3) were tested in AoU and GLAD (individual-level data access), using the same models, and meta-analyzed as above. Each of the 17 PGS other than depression was tested unadjusted and adjusted for the depression PGS (35 tests), with FDR correction applied jointly given the correlation among PGS and models. To identify which components of educational attainment (EA) drive its association, its cognitive (Cog) and non-cognitive (NonCog) components (35) were also tested.

### 2.6 Pathway, transcriptomic and drug-target analyses

#### Serotonin-Related and all KEGG Pathway Analyses

To extend the FUMA gene-set analysis to all current KEGG pathways, we ran gene-set analyses in MAGMA (v1.10) (26) using the current KEGG release. We tested gene-set enrichment in four pre-specified serotonin-related KEGG pathways (serotonergic synapse, neuroactive ligand signaling, tryptophan metabolism, and folate biosynthesis) (36,37), and all 369 KEGG pathways. FDR correction was applied separately to each set, and gene-level p-values were examined for enriched pathways to identify signal drivers.

#### Transcriptome- and proteome-wide association analyses (TWAS and PWAS)

To test whether self-reported SSRI response is associated with genetically predicted gene expression or protein abundance, we performed transcriptome- and proteome-wide association analyses (TWAS and PWAS) using FUSION (38) and summary-based Mendelian randomization (SMR) (39), with brain and blood expression and protein quantitative trait locus (eQTL, pQTL) panels. FDR correction was applied across panels within each method, separately for TWAS and PWAS.

#### Prioritizing convergent candidate genes

Genes were prioritized as convergent candidates if they reached p <0.01 in the MAGMA gene-based analysis, and in at least one brain panel for both TWAS and PWAS (Supplementary Note5). These thresholds were used for prioritization, not as tests of significance.

#### Drug-target enrichment

To aid clinical interpretation, we tested whether gene-level associations were enriched for known drug targets using MAGMA (26) and TWAS-based Gene Set Enrichment Analysis (TWAS-GSEA; (40)), with drug-gene interactions from DrugTargetor (41). TWAS-GSEA is directional, testing whether a drug’s predicted effect on expression aligns with or opposes the expression profile associated with response. Drug-level results were FDR-corrected. At the Anatomical Therapeutic Chemical (ATC) class level, for classes with at least five drugs, signed TWAS-GSEA statistics within each class were compared with those of all other drugs (two-sided Wilcoxon rank-sum test; FDR-corrected across classes). As drugs within a class often share targets, class-level results should be interpreted cautiously.

## 3. Results

### 3.1 Study overview

After quality control, 62,879 participants of European-like genetic ancestry were retained across seven cohorts, of whom 45,642 (73%) individuals were self-reported SSRI responders and 17,237 (27%) were non-responders, with response proportions ranging from 66% to 82% across cohorts (Table1).

**Table 1.**
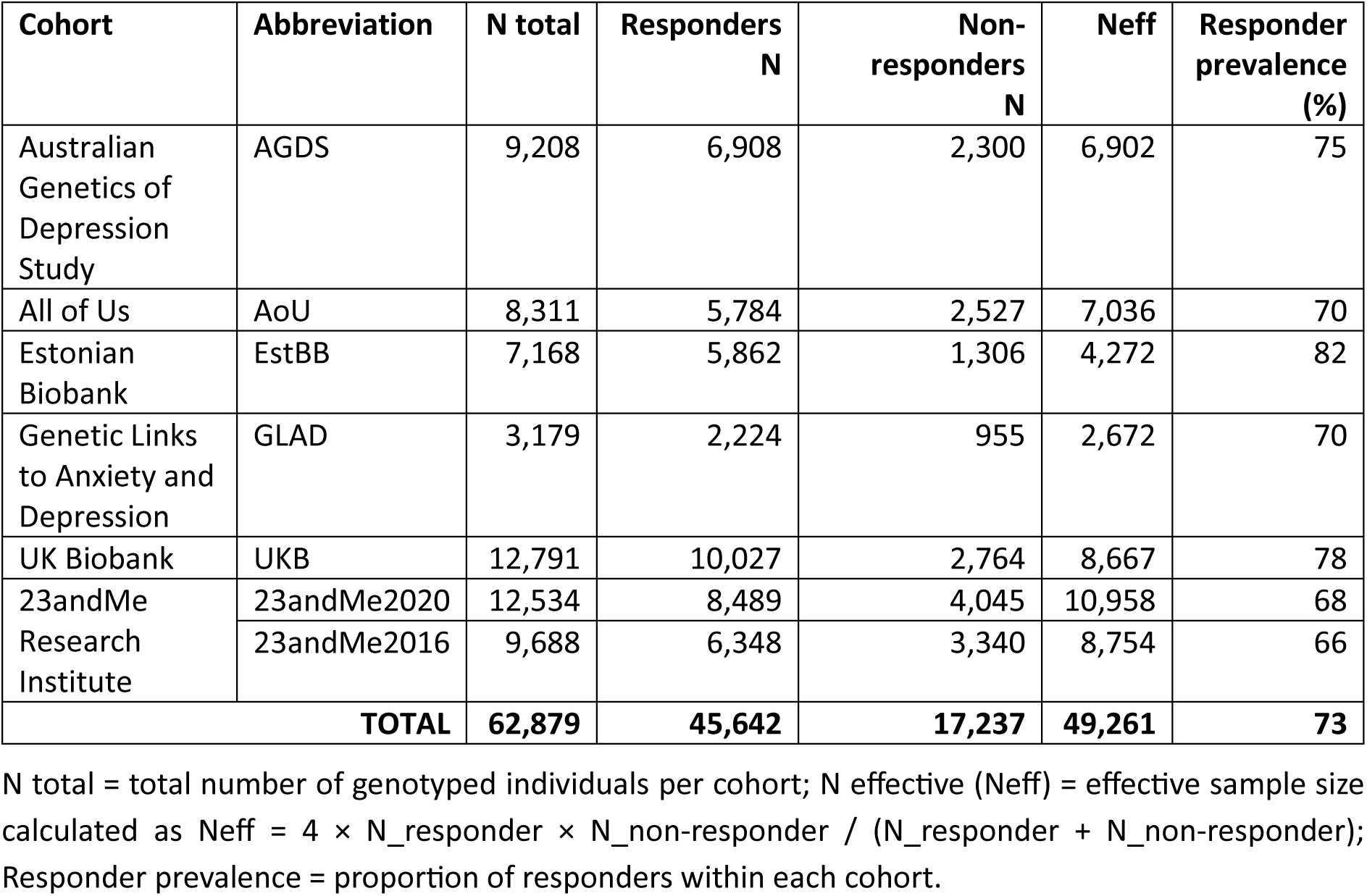
Cohort-level sample sizes and effective sample sizes (Neff) for self-reported SSRI response across seven contributing cohorts.

### 3.2 Genome-wide meta-analysis

The primary fixed-effect (FE) meta-analysis (Neff = 49,261; 9,033,479 variants) identified no genome-wide significant SNPs (Fig.1), 14 independent loci reached suggestive significance (p <5×10⁻^6^) (Supplementary TableS4). Genomic inflation was negligible (λ = 1.02) (Supplementary Fig.S1), and heterogeneity across cohorts was modest overall (mean I² = 13.4%; 5.4% of SNPs nominally heterogeneous, p_Q < 0.05).

**Figure 1:**
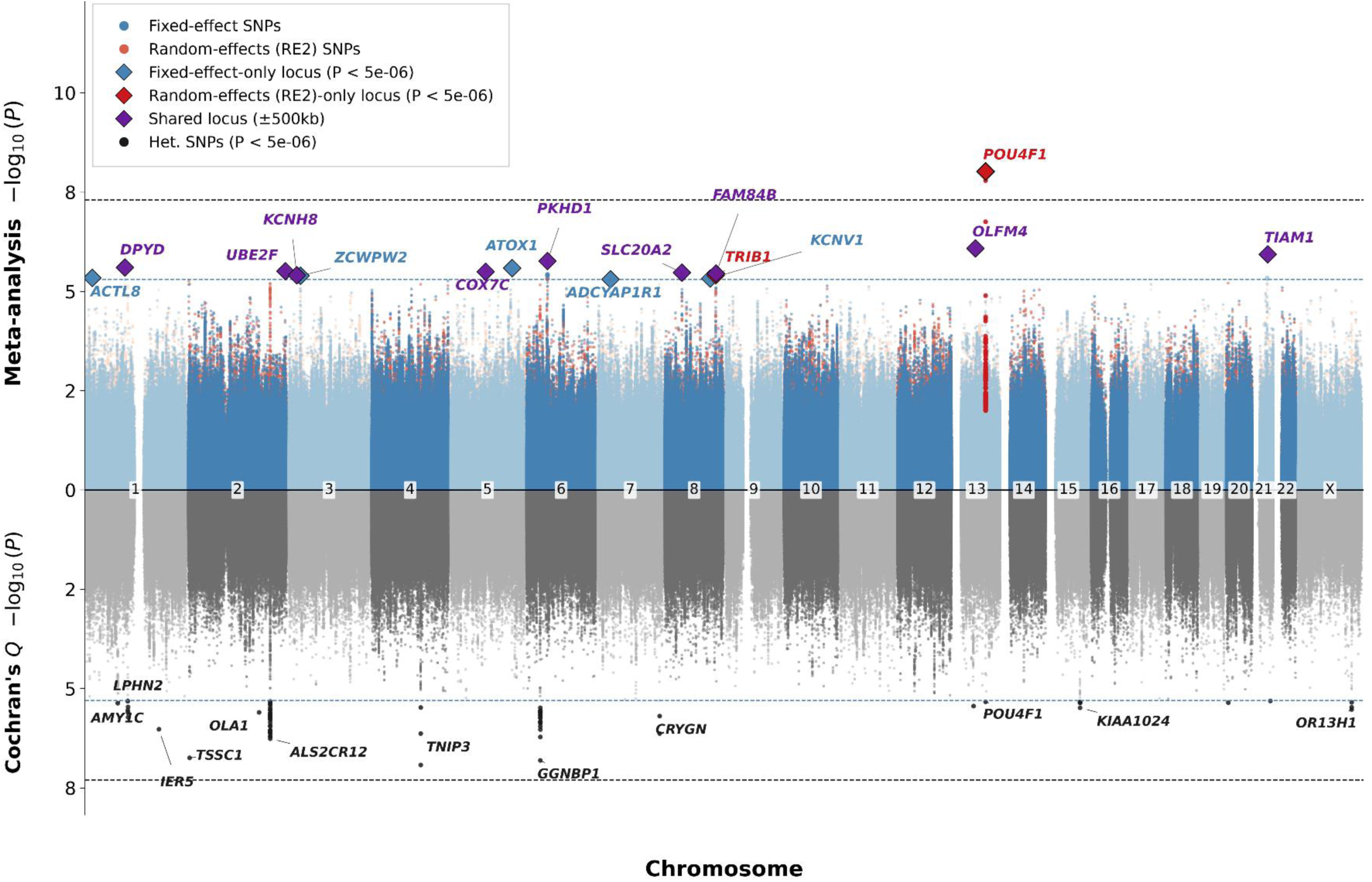
GWAS meta-analysis of self-reported SSRI response, with Fixed Effect (FE) vs Random Effect (RE2) meta-analysis and Cochran’s. **Q.** Miami plot of the self-reported SSRI response meta-analysis (45,642 responders, 17,237 non-responders; N = 62,879). Top: −log10(P) from the fixed-effect inverse-variance-weighted (FE) and random-effects (RE2) models. Bottom: −log10(P) for Cochran’s Q test of heterogeneity. X-axis: chromosomal position (chromosomes 1–22 and X). Dashed black line: genome-wide significance (P = 5×10⁻⁸); blue line: suggestive threshold (P = 5×10⁻⁶). Labels show the nearest protein-coding gene for SNPs with P < 5×10⁻⁶

Under the Han-Eskin random-effects model (RE2; Supplementary TableS5), one locus reached genome-wide significance: rs4884091 at 13q22.3 (p = 9.48×10⁻⁹). Associations at this locus were heterogeneous across cohorts (I² = 84%; p_Q = 5.31×10⁻⁶), and it was not significant under FE (p = 5.00×10⁻⁵) or conventional random effects (p = 0.17). The signal was driven by 23andMe 2020 (m = 1.00, the posterior probability of an effect in that cohort; p = 2.09 × 10⁻⁸), with weaker support from other cohorts, while UKB showed an opposite direction of effect (Supplementary Fig.S2). Figure 2 shows a proxy, rs4278634 (1.1kb away; LD-r^2^ = 0.84 in 1KGP3 EUR ((42))), as the lead SNP was not available in GLAD which showed similar heterogeneity (I² = 81%), and UKB discordance. By contrast, the top FE locus, rs73200007 (p = 8.25×10⁻⁷), showed no heterogeneity (I² = 0%; p_Q = 0.78; Supplementary Fig.S2, Fig.S3).

**Figure 2:**
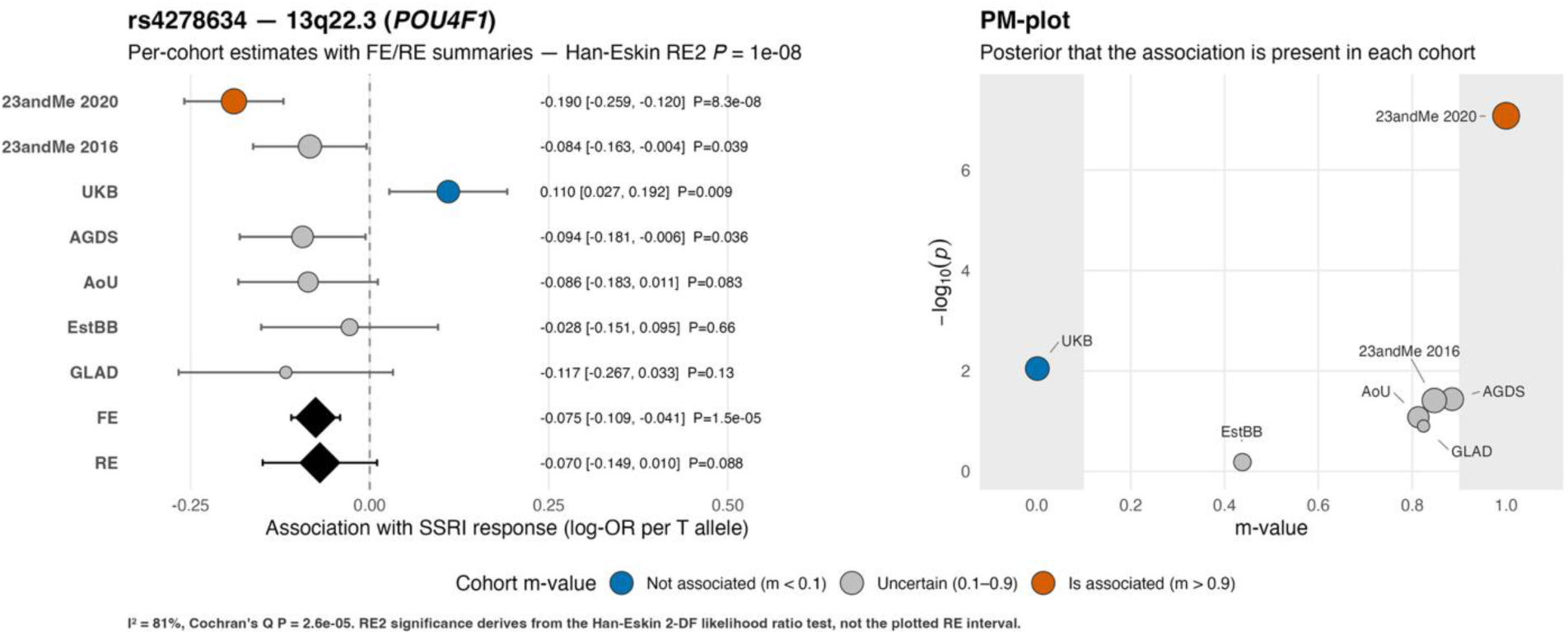
Between-cohort heterogeneity at locus rs4278634 (13q22.3, near *POU4F1*). Per-cohort associations at rs4278634 (13q22.3; nearest gene POU4F1), a proxy for the RE2 lead SNP rs4884091, which was not available in GLAD. **(A)** Forest plot of per-cohort log-OR (odds ratio) of self-reported SSRI response per T allele, with 95% confidence intervals, and the fixed-effect (FE) and conventional random-effects (RE) summary estimates (diamonds). Point color indicates each cohort’s m-value, the posterior probability that an effect is present: not associated (m < 0.1), uncertain (0.1–0.9) or associated (m > 0.9). Effects were heterogeneous across cohorts (I² = 81%; Cochran’s Q p = 2.6 × 10⁻⁵). The RE2 p-value (1 × 10⁻⁸) comes from a two-degree-of-freedom likelihood ratio test and is not represented by the plotted RE interval. **(B)** PM-plot of each cohort’s −log₁₀(p) against its m-value. Shaded regions mark m < 0.1 and m > 0.9. The top FE locus (rs73200007), which showed no heterogeneity, is shown in Supplementary Fig.S3.

### 3.3 Characterization and functional annotation of GWAS SNPs

FUMA defined 14 independent suggestive loci, each tagged by a single lead SNP. Positional mapping identified seven protein-coding genes across five of these loci: *SCLY, TIAM1, SLC20A2, PKHD1, KCNH8, UBE2F, and UBE2F-SCLY* (Supplementary TableS6). Four loci had prior GWAS Catalog associations (release e0_r2022-11-29), mostly with endocrine and renal traits such as sex hormone-binding globulin and glomerular filtration rate. The neurobehavioral overlaps were with insomnia and Parkinson’s disease (Supplementary TableS7).

No gene reached significance in the MAGMA gene-based analysis (20,068 genes; Bonferroni p <2.49×10⁻⁶). The strongest signal was for *TIAM1* (p = 9.7×10⁻⁶), also the positionally mapped gene at locus 14 (Supplementary TableS10). Gene-set, tissue-expression and GENE2FUNC analyses showed no significant enrichment, although brain tissues ranked highest amongst the 54 specific tissues (9 of the top 11; Supplementary Figs.S5-S8).

Applying the same pipeline to the RE2 summary statistics identified 11 suggestive loci (p <5×10⁻⁶), 9 overlapping the FE set and 2 specific to RE2, including the genome-wide significant rs4884091 locus (Supplementary TableS5). Positional mapping implicated the same seven genes, and there was no significant gene-set or tissue-expression enrichment.

### 3.4 SNP-based heritability estimation and genetic correlations

Self-reported SSRI response showed modest but significant SNP-based heritability (LDSC; *h²* = 3.1%, SE = 1.4%; p = 0.030), with an LDSC intercept of 1.012 (SE = 0.006) not significantly different from 1, indicating minimal residual population stratification.

Six of the 15 comparator traits showed significant genetic correlations with self-reported SSRI response after FDR correction (q<0.05; Table2). Negative correlations were seen with PTSD (*rg* =-0.83, q =0.004), ADHD (*rg* =-0.78, q =0.012), depression (rg =-0.73, q =0.011), anxiety (*rg* =-0.59, q =0.003), and BMI (*rg* =-0.25, q =0.029) and a positive genetic correlation with educational attainment (*rg* =0.49, q =0.006). Given the modest *SNP-h²* of self-reported SSRI response (3.1%), confidence intervals were wide, meaning only traits with substantial genetic overlap could be detected. Two antidepressant treatment-response phenotypes were excluded because their *SNP-h²* estimates were non-significant (non-remission) or negative (percentage improvement) (Supplementary TableS8).

**Table 2.**
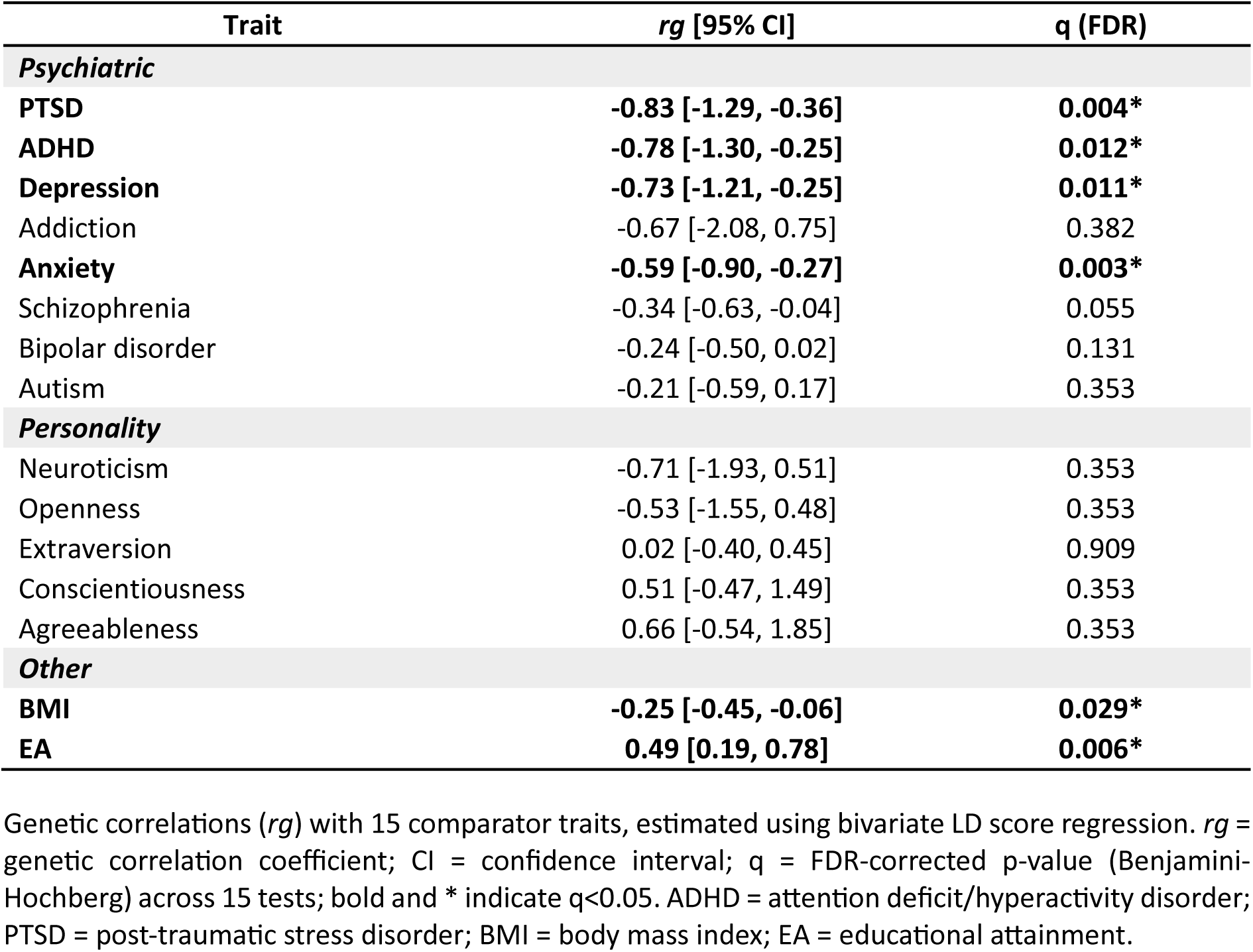
Genetic correlations between self-reported SSRI response and comparator traits.

| **Trait** | ***rg* [95% CI]** | **q (FDR)** |
|---|---|---|
| ***Psychiatric*** |  |  |
| **PTSD** | **-0.83 [-1.29, -0.36]** | **0.004\*** |
| **ADHD** | **-0.78 [-1.30, -0.25]** | **0.012\*** |
| **Depression** | **-0.73 [-1.21, -0.25]** | **0.011\*** |
| Addiction | -0.67 [-2.08, 0.75] | 0.382 |
| **Anxiety** | **-0.59 [-0.90, -0.27]** | **0.003\*** |
| Schizophrenia | -0.34 [-0.63, -0.04] | 0.055 |
| Bipolar disorder | -0.24 [-0.50, 0.02] | 0.131 |
| Autism | -0.21 [-0.59, 0.17] | 0.353 |
| ***Personality*** |  |  |
| Neuroticism | -0.71 [-1.93, 0.51] | 0.353 |
| Openness | -0.53 [-1.55, 0.48] | 0.353 |
| Extraversion | 0.02 [-0.40, 0.45] | 0.909 |
| Conscientiousness | 0.51 [-0.47, 1.49] | 0.353 |
| Agreeableness | 0.66 [-0.54, 1.85] | 0.353 |
| ***Other*** |  |  |
| **BMI** | **-0.25 [-0.45, -0.06]** | **0.029\*** |
| **EA** | **0.49 [0.19, 0.78]** | **0.006\*** |

Genetic correlations (*rg*) with 15 comparator traits, estimated using bivariate LD score regression. *rg* = genetic correlation coefficient; CI = confidence interval; q = FDR-corrected p-value (Benjamini-Hochberg) across 15 tests; bold and \* indicate q<0.05. ADHD = attention deficit/hyperactivity disorder; PTSD = post-traumatic stress disorder; BMI = body mass index; EA = educational attainment.

### 3.5 Polygenic scoring analyses

#### Shared genetic architecture with psychiatric and other traits

PGS analyses in AoU and GLAD were combined by fixed-effect meta-analysis (Fig.3, Supplementary TablesS9). All psychiatric trait PGS, except bipolar disorder, were FDR-significant and similar in effect size: depression (OR = 0.86, p = 1.6×10⁻^12^), ADHD (OR = 0.88, p = 6.4×10⁻^10^), PTSD (OR = 0.88, p = 3.2×10⁻^9^), anxiety (OR = 0.89, p = 6.1×10⁻^8^), addiction (OR = 0.92, p = 6.6×10⁻⁵), autism (OR = 0.93, p = 5.4×10⁻⁴) and schizophrenia (OR = 0.94, p = 3.9×10⁻³). Cohort-specific associations are in Supplementary TableS9.

**Figure 3.**
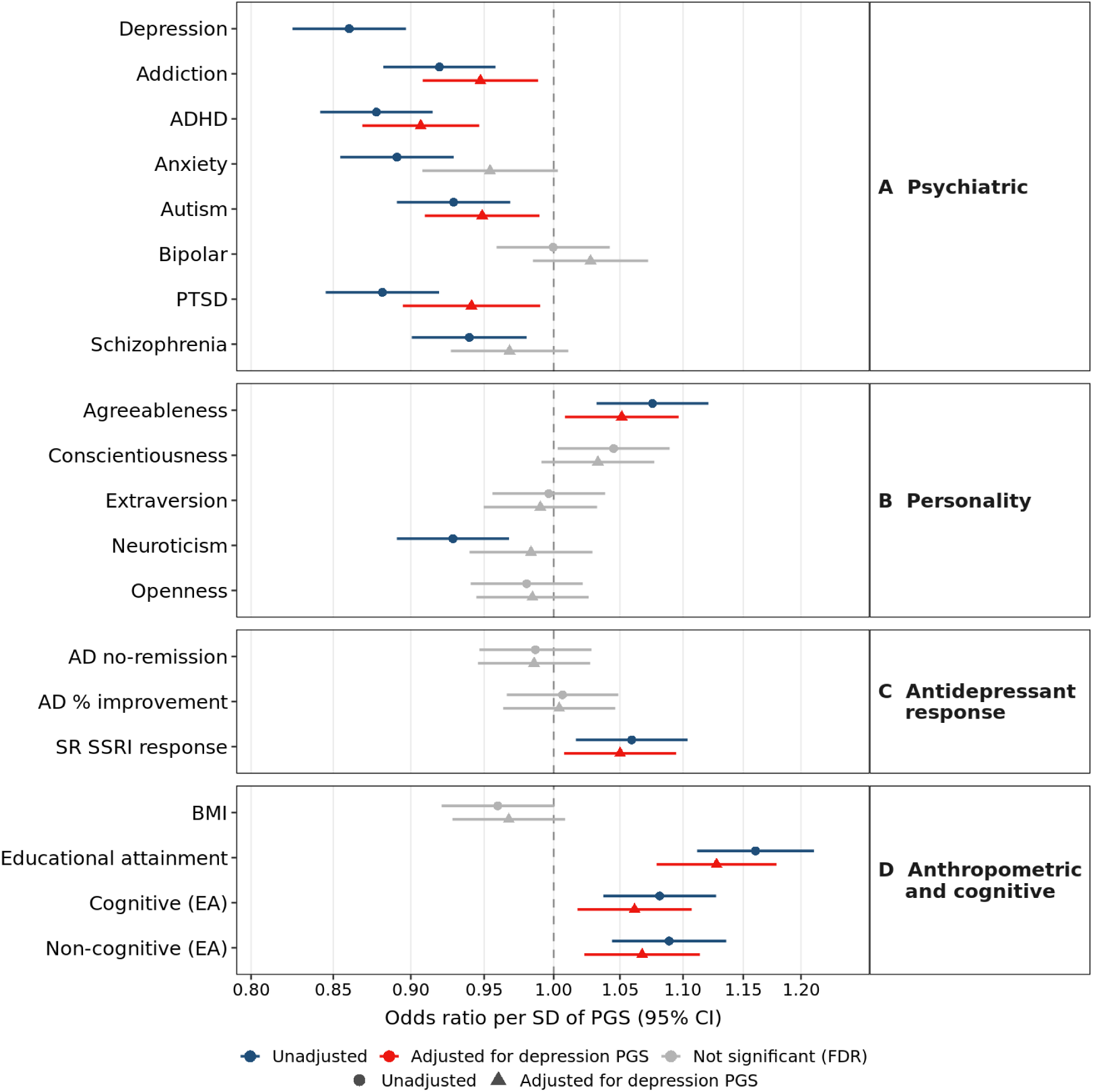
Polygenic score associations with self-reported SSRI response. Odds ratios (95% CI) per SD of SBayesRC polygenic score (PGS), from fixed-effect meta-analysis of GLAD and AoU, for (A) psychiatric, (B) personality, (C) antidepressant response and (D) anthropometric and cognitive traits. Circles show unadjusted models and triangles models adjusted for depression PGS; the depression PGS was tested unadjusted only. Blue (unadjusted) and red (adjusted) indicate FDR significance (q < 0.05) across 35 tests; gray indicates non-significance. FDR correction for the cognitive (Cog) and non-cognitive (NonCog) components of EA were applied within the EA decomposition (EA, Cog and NonCog). Cohort-specific results are in Supplementary Table S9. AD = antidepressant; ADHD = attention-deficit/hyperactivity disorder; BMI = body mass index; EA = educational attainment; PTSD = post-traumatic stress disorder; SR = self-reported.

Among personality traits, higher agreeableness PGS was associated with increased odds of response (OR = 1.08, p = 5.4×10⁻⁴) and higher neuroticism PGS with reduced odds of response (OR = 0.93, p = 4.3×10⁻⁴), both FDR-significant. EA PGS showed one of the strongest associations, with higher EA PGS associated with increased odds of response (OR = 1.16, p = 1.3×10^-11^), comparable in magnitude to depression PGS. Both the cognitive (Cog; OR = 1.08, p = 2.3×10⁻^4^) and non-cognitive (NonCog; OR = 1.09, p = 7.6×10⁻^5^) components of EA were associated with increased odds of response, with similar effect sizes. The leave-one-out (LOO) SSRI-response PGS was also associated with increased odds of response (OR = 1.06, p = 6.1×10⁻³).

To assess whether these associations were independent of shared genetic liability to depression, we repeated the analysis adjusting for depression PGS. Traits remaining significant after adjustment were EA (OR = 1.13, p = 1.0×10⁻^7^), ADHD (OR = 0.91, p = 8.3×10⁻^6^), addiction (OR = 0.95, p = 0.013), autism (OR = 0.95, p = 0.015), agreeableness (OR = 1.05, p = 0.019) and PTSD (OR = 0.94, p = 0.019), and the LOO SSRI-response PGS (OR = 1.05, p = 0.020), as well as both EA components (Cog: OR = 1.06, p = 5.5×10⁻³; NonCog: OR = 1.07, p = 2.7×10⁻³).

#### Leave-one-out polygenic scoring prediction

LOO PGS for self-reported SSRI response, derived by excluding each target cohort from the discovery meta-analysis, showed directionally consistent associations with self-reported SSRI response in all three target cohorts (Fig.4): (AGDS: incremental liability-scale R² = 0.05%, OR = 1.04, 95% CI: 0.99–1.09, p = 0.133, AoU: R² = 0.08%, OR = 1.06, 95% CI: 1.01–1.11, p = 0.026; GLAD: R² = 0.14%, OR = 1.06, 95% CI: 0.99–1.15, p = 0.110). There was no evidence of heterogeneity across cohorts (Cochran’s Q = 0.44, p = 0.803, *I²* = 0%). The pooled FE meta-analysis was significant (OR = 1.05, 95% CI: 1.02–1.08, p = 0.002).

**Figure 4.**
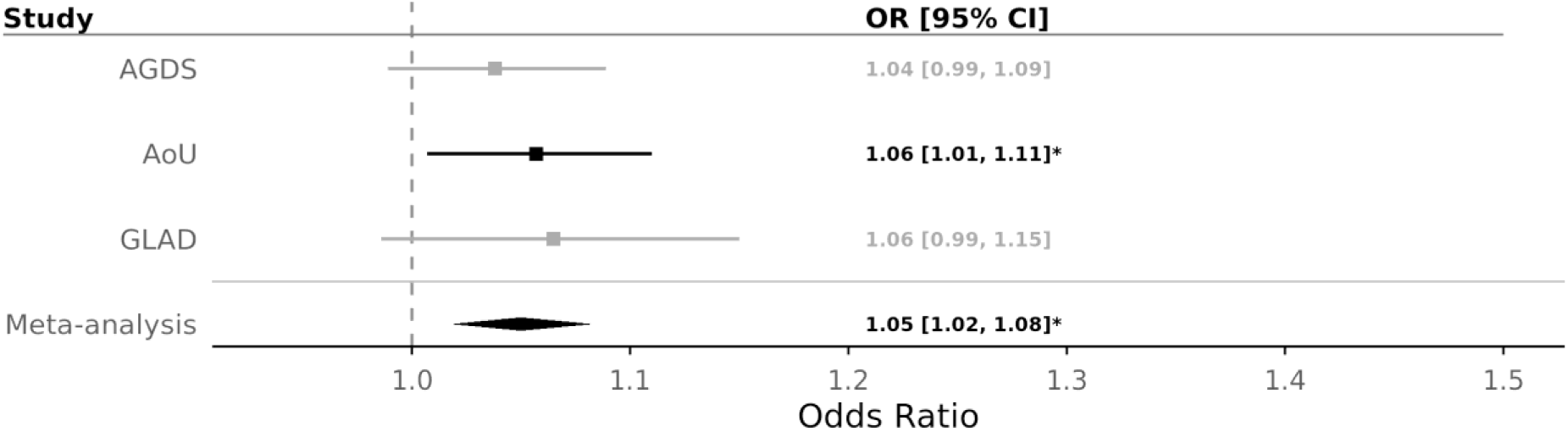
Association between the leave-one-out SSRI-response polygenic score (PGS) and self-reported SSRI response in the Australian Genetics of Depression Study (AGDS), All of Us (AoU) and the Genetic Links to Anxiety and Depression study (GLAD), and their fixed-effect meta-analysis. Each cohort’s PGS was derived from a discovery meta-analysis excluding that cohort. Squares show cohort-specific odds ratios (OR) per SD of the PGS with 95% confidence intervals; the diamond shows the pooled fixed-effect estimate, with its width spanning the 95% confidence interval. The dashed vertical line indicates OR = 1. Sample sizes (total N; responders/non-responders) were: GLAD, N = 3,121 (2,186/935); AoU, N = 7,904 (5,501/2,403); AGDS, N = 8,940 (6,697/2,243); meta-analysis, N = 19,965 (14,384/5,581). Non-significant estimates (p ≥ 0.05) are shown in grey; *p < 0.05 (nominal).

### 3.6 Serotonergic and KEGG pathway enrichment

None of the four pre-specified serotonin-related KEGG pathways were enriched (all q≥0.29; Supplementary TableS11). Across all 369 KEGG pathways, the nicotine addiction pathway (hsa05033; 39 genes) was significantly enriched (all-pathway q=0.029; p = 8.0×10⁻⁵, Supplementary TableS12). The signal was distributed across eight modestly associated genes rather than driven by a single gene (Supplementary TableS13): GABA-A receptor subunit genes (*GABRG1*, p = 0.006; *GABRA2*, p = 0.013; *GABRR3*, p = 0.037; *GABRG3*, p = 0.045; *GABRR1*, p = 0.048), the vesicular GABA transporter *SLC32A1* (p = 0.036), the AMPA receptor subunit *GRIA4* (p = 0.013) and the vesicular glutamate transporter *SLC17A6* (p = 0.005). No other pathway survived correction; the next strongest were long-term depression (synaptic plasticity pathway; hsa04730) and Alzheimer’s disease (both p = 0.004, q=0.45).

### 3.7 Transcriptome- and proteome-wide association

No TWAS or PWAS association survived FDR correction with either FUSION or SMR (Supplementary TablesS14–S17). Combining gene-level (MAGMA), transcriptomic and proteomic evidence in brain panels, six genes reached p <0.01 across all three lines: *ISLR, SLC17A6, DHRS7, FUT8, GSTZ1* and *HADH* (Table3; Supplementary TableS18). A further 67 genes were supported by two lines (Supplementary TableS19).

**Table 3.** Convergent-evidence candidate genes for SSRI response.

| Gene | MAGMA |  |  | TWAS |  |  | PWAS |  |  | Concordant evidence |  |
|---|---|---|---|---|---|---|---|---|---|---|---|
|  | *Best P* | *P<0.01* | *P<0.001* | *Best P* | *P<0.01* | *P<0.001* | *Best P* | *P<0.01* | *P<0.001* | *P<0.01* | *P<0.001* |
| *ISLR* | 2.43E-03 | ✓ |  | 4.28E-03 | ✓ |  | 6.81E-04 | ✓ | ✓ | 3 | 1 |
| *SLC17A6* | 4.62E-04 | ✓ | ✓ | 7.91E-03 | ✓ |  | 4.09E-03 | ✓ |  | 3 | 1 |
| *DHRS7* | 3.97E-03 | ✓ |  | 2.06E-03 | ✓ |  | 2.16E-03 | ✓ |  | 3 | 0 |
| *FUT8* | 3.25E-03 | ✓ |  | 2.35E-03 | ✓ |  | 3.06E-03 | ✓ |  | 3 | 0 |
| *HADH* | 6.24E-03 | ✓ |  | 7.67E-03 | ✓ |  | 6.64E-03 | ✓ |  | 3 | 0 |
| *GSTZ1* | 6.69E-03 | ✓ |  | 6.00E-03 | ✓ |  | 7.52E-03 | ✓ |  | 3 | 0 |

*Genes prioritised by a convergent-evidence framework requiring nominal support (p < 0.01) across three independent lines: positional (MAGMA gene-based association or nearest gene to a lead variant), transcriptomic (TWAS by FUSION or SMR) and proteomic (PWAS by FUSION or SMR). The six genes shown met all three lines at p < 0.01. For each line, the strongest (minimum) p-value across constituent tests is given, with support marked at p < 0.01 and at the stricter p < 0.001 threshold. “Concordant evidence: 3 lines” columns count the lines meeting each threshold. No association survived genome-wide multiple-testing correction; these candidates rest on sub-threshold signal and require replication. MAGMA, Multi-marker Analysis of GenoMic Annotation; TWAS, transcriptome-wide association study; PWAS, proteome-wide association study; SMR, summary-based Mendelian randomisation; Y = Yes, meets criteria.*

### 3.8 Drug-target enrichment

Drug-gene interactions from Drug Targetor (41) were used to test for drug-target enrichment with MAGMA and TWAS-GSEA.

MAGMA tested enrichment across 111 ATC classes and 1,454 individual compounds (Supplementary TableS20-S22). Three ATC classes showed significant enrichment: hypnotics and sedatives (N05C; q=4.4×10⁻^10^), anxiolytics (N05B; q=1.4×10⁻³) and general anesthetics (N01A; q=3.0×10⁻^2^) (Fig.5A). As drugs in these classes largely share GABA-A receptor targets, these class-level tests are not independent and should be interpreted cautiously. No individual compound survived correction (min q=0.42).

**Figure 5:**
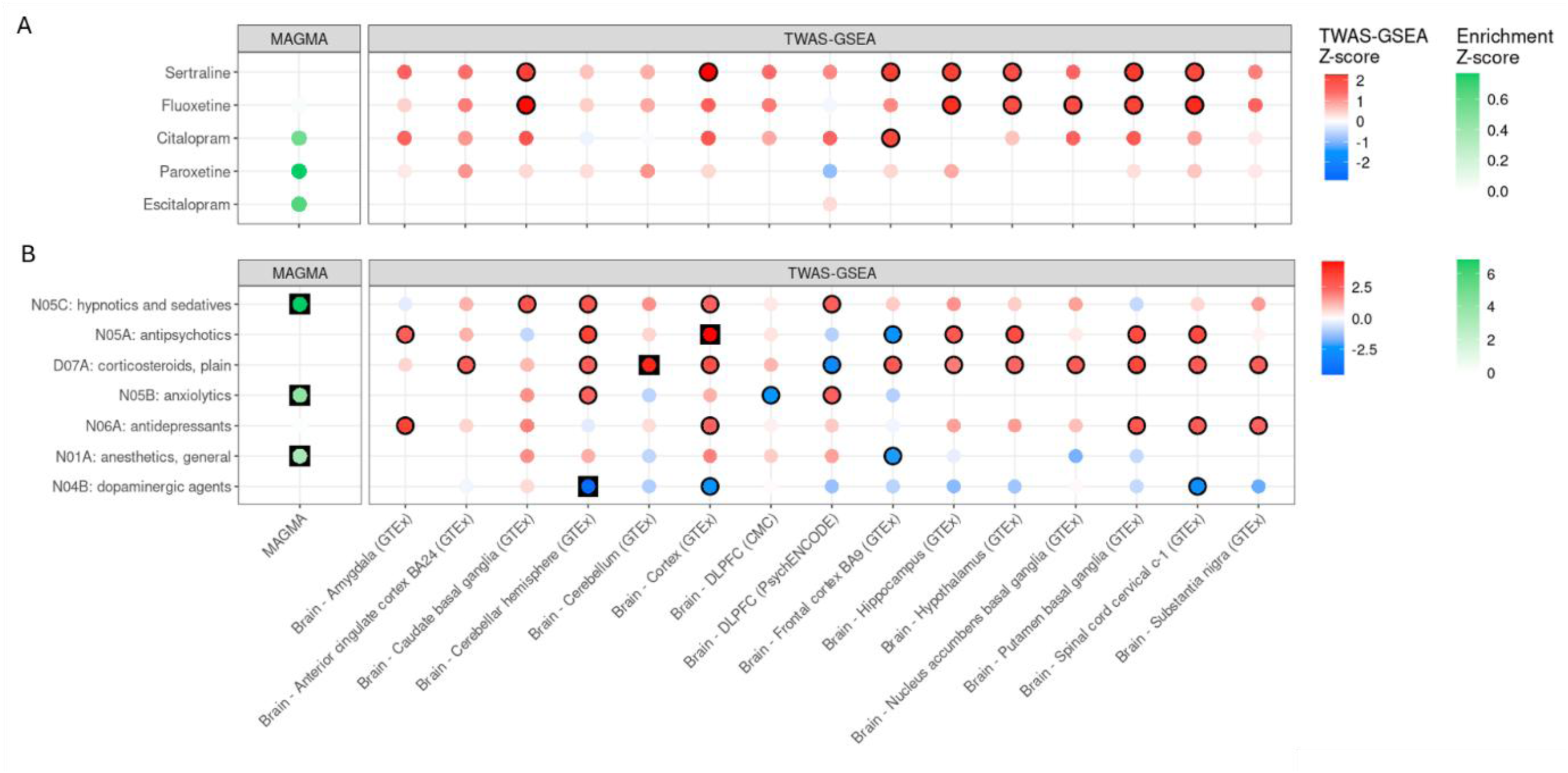
Drug Enrichment at the Anatomical Therapeutic Chemical (ATC) level and for selected Selective serotonin reuptake inhibitors (SSRIs) (A) SSRI enrichment Z-score per method/expression-panel combination. (B) ATC-class enrichment Z-score per method/expression-panel combination. TWAS-GSEA uses a diverging red–blue palette: red indicates drugs (or ATC classes) whose known effects on gene expression oppose the expression profile genetically predicted in responders; blue indicates effects in the same direction. MAGMA uses a sequential white-to-green palette: green indicates enrichment for the trait association signal (no directional interpretation). Black circles: nominally significant (p < 0.05); black squares: FDR-significant (q < 0.05).

TWAS-GSEA tested whether the direction of predicted transcriptional effects of individual SSRIs and drug classes were concordant or discordant with the direction of the genetic signature of SSRI response (Supplementary TableS21-23). Sertraline (seven brain tissues), fluoxetine (six) and citalopram (one) reached nominal significance, each inducing expression signatures opposing the SSRI-responder signature, but none survived correction (Fig. 5A, Supplementary TableS21). Across 115 ATC classes, three drug-class-tissue combinations were FDR-significant (Fig.5B, Supplementary TableS23): dopaminergic agents (N04B; cerebellar hemisphere; q=0.01) were concordant with the transcriptional direction of the SSRI-responder expression signature, while antipsychotics (N05A; cortex; q=7.6×10⁻³) and corticosteroids (D07A; cerebellum; q=1.5×10⁻²) showed opposing transcriptional direction. Antidepressants (N06A) reached nominal significance in five brain tissues, also opposing, consistent with drug-level results. No class or compound showed convergent evidence across MAGMA and TWAS-GSEA.

## 4. Discussion

In the largest GWAS meta-analysis restricted to self-reported SSRI response, we found modest but significant SNP-based heritability (3.1%), with diagnostics indicating a polygenic signal rather than artefact. However, no locus reached genome-wide significance in the primary fixed-effect analysis, and gene-based analysis was similarly null, with *TIAM1* the strongest sub-threshold signal. At the current sample size and heritability, this GWAS appears underpowered for common-variant discovery; larger samples or more precise phenotyping will be needed.

Our work differs from a recent GWAS by Koch et al. (8) in that we used a single, consistently ascertained phenotype (self-reported response), restricted analyses to SSRIs and, where possible, to individuals with lifetime MDD, and added a new cohort, All of Us. Koch et al. combined self-reported, clinician-rated and registry-based outcomes (N ≈135,000), identifying two novel loci for non-response (one SSRI-specific); their self-reported subset (N ≈60,000) overlaps our cohorts, precluding its inclusion here.

In our Han-Eskin random-effects sensitivity analysis, one locus (rs4884091) reached genome-wide significance, driven by 23andMe2020. This locus did not overlap Koch et al.’s genome-wide significant locus but corresponded to their strongest self-report-restricted signal, which fell just below significance in their full analysis. This likely reflects the same 23andMe-derived association, reported by Li et al. (20), resurfacing in each analysis rather than three independent findings. The lead SNP at our *TIAM1* locus, rs6517046, also reached suggestive significance in their full and self-report-restricted analyses, with a concordant direction of effect (8).

*TIAM1* encodes a Rac1 guanine nucleotide exchange factor that acts at excitatory synapses and is implicated in neuronal development (43). In mouse models of chronic pain, Tiam1 stabilizes synaptic N-methyl-D-aspartate (NMDA) receptors in the anterior cingulate cortex (ACC), driving structural plasticity associated with ACC hyperactivity and depressive-like behavior, which low-dose ketamine reverses by blocking this pathway (44). This places *TIAM1* in a plausibly relevant glutamatergic pathway, though the evidence concerns ketamine, not SSRIs, and the finding warrants replication.

SSRIs block serotonin reuptake within hours, yet clinical improvement typically takes weeks, suggesting additional mechanisms, such as neuroplastic adaptation, are required for response (45). We found no enrichment for serotonergic pathway genes, although this null may reflect limited power in small gene sets. Instead, the FDR-significant KEGG nicotine addiction pathway was driven by GABA-A receptor and transporter genes, the glutamate transporter gene *SLC17A6,* and an AMPA receptor gene (*GRIA4*), with no contribution from nicotinic receptor genes, suggesting GABAergic and glutamatergic rather than nicotine-specific biology. Drug-target enrichment for hypnotics, anxiolytics and general anesthetics, driven largely by anesthetics rather than ketamine, likewise points to GABA-A modulation. GABAergic deficits are well documented in MDD (46,47), glutamatergic and NMDA receptor dysfunction is implicated in MDD neuroimaging (48), and SSRIs also influence GABAergic neurotransmission (49). *SLC17A6* showed sub-threshold evidence across gene-level, pathway, transcriptome and proteome analyses, and *TIAM1*’s role in synaptic plasticity offers a further route to downstream circuit change. As these pathways are not SSRI-specific and power may differ across gene sets, we interpret them cautiously. Self-reported response, unconstrained by a fixed assessment window, may capture delayed adaptation, plausibly contributing to the higher rates of self-reported benefit and the non-serotonergic signals observed here.

In TWAS-GSEA, where direction was interpretable, citalopram, fluoxetine and sertraline showed nominal expression effects opposing the responder-associated profile, as did antipsychotics and corticosteroids (FDR-significant), whereas dopamine-enhancing agents aligned with it. Two non-exclusive interpretations are possible. Opposition suggests a drug counteracting an expression state that predisposes to poorer outcomes, so responders are those whose baseline profile is most amenable to reversal along that drug’s axis of action. Alignment suggests a drug shifting expression towards a profile responders already carry, so reproducing rather than reversing it. Both could operate for different drug classes, and the present data cannot distinguish them; as the responder profile reflects genetically predicted expression rather than measured pre- and post-treatment expression, these results are hypothesis-generating.

The leave-one-out self-reported SSRI-response PGS was significantly associated with self-reported response, indicating the GWAS captures genuine common-variant signal despite insufficient power for locus discovery, as in other psychiatric traits where SNP-based heritability is detectable well before genome-wide significant loci emerge (50,51). Genetic liability to most psychiatric disorders was associated with poorer self-reported response, extending beyond shared liability to depression for ADHD, PTSD, autism and addiction, but not for anxiety and schizophrenia. The persistent ADHD association is consistent with prior work linking ADHD PGS to treatment-resistant depression (52). Most associations were supported by genetic correlations; those significant only in PGS analyses (autism, addiction and schizophrenia) likely reflect the lower power of *rg* estimation. These associations may reflect biological non-response in individuals with greater comorbid psychiatric burden, behavioral factors linked to these traits, or misdiagnosis, where depression secondary to an unrecognized condition such as ADHD is treated as MDD, so that apparent non-response reflects an unaddressed diagnosis rather than pharmacological failure (53). Although these PGS, like the SSRI-response PGS, explain too little variance to guide individual treatment, if replicated, polygenic loading for comorbid traits could help identify patients who warrant closer monitoring or diagnostic reassessment.

Among personality traits, only agreeableness remained associated with better response after adjustment for depression PGS, whereas the neuroticism association largely reflected shared liability to depression. Agreeableness has been linked to stronger therapeutic alliance and modestly better outcomes in psychotherapy and pharmacotherapy (54), but also to an approval-oriented, socially desirable self-presentation (55), which may inflate self-reported treatment benefit.

EA PGS was among the strongest and most robust predictors of better self-reported response. It captures genetic influences on cognitive ability, socioeconomic status and health-related behaviors (56,57), and can be decomposed into a cognitive (Cog) component, shared with cognitive performance, and a non-cognitive (NonCog) component, linked to personality, decision-making and lower health-risk behavior (35). Both components were associated with better response, with similar effect sizes, independent of depression PGS. As both are similarly genetically correlated with income (35), socioeconomic factors are a plausible shared contributor. This is consistent with evidence that EA and cognitive performance PGS distinguish treatment-resistant from treatment-responsive depression in electronic health records (58).

Several pathways could link these components to response. Antidepressant treatment requires patients to persist through early side effects and delayed benefit, and to recognize when to seek review. Cognitive ability may support such persistence, consistent with links between health literacy and adherence (59). Non-cognitive traits such as persistence, future orientation and engagement with healthcare may similarly support adherence and follow-up, while socioeconomic advantage, captured by both components, may improve access to and continuity of care. These pathways were not testable here, and the associations may also reflect how response is reported.

This study has several limitations. Our phenotype relies on cross-sectional, retrospective self-report, which is subject to recall and reporting bias and may capture improvements in well-being not specific to antidepressant pharmacology, such as changing life circumstances, placebo response or non-specific therapeutic benefit. Question wording and response options differed across cohorts, which may have reduced power for an already low-heritability trait. Genetic contributions to response may also change over the lifespan, warranting longitudinal data. Available data remains overwhelmingly of European ancestry, reflecting the underrepresentation of non-European populations in genetic and mental health research, compounded by underrepresentation in follow-up questionnaires from which response phenotypes were derived. Addressing these gaps will require larger, multi-ancestry samples; functional follow-up of the GABAergic and glutamatergic signals, particularly *SLC17A6*; and work disentangling general psychiatric liability from treatment-response-specific genetics.

In conclusion, self-reported SSRI response captures genuine genetic signal relevant to antidepressant response, although current samples remain underpowered for common-variant discovery. Our findings implicate GABAergic and glutamatergic mechanisms alongside broader psychiatric burden: higher genetic liability for ADHD, PTSD, autism and addiction was associated with poorer self-reported response independent of depression liability, and genetic propensity for higher educational attainment with better response. As a patient-centered and scalable phenotype, self-report offers a practical route to the larger samples this trait requires.

## Data Availability

The GWAS summary statistics and polygenic scores produced in this study (excluding data from 23andMe Research Institute) will be deposited in the GWAS Catalog and PGS Catalog on publication. WGS data are available via the UKB research analysis platform (https://ukbiobank.dnanexus.com/landing), which is open to researchers from academic, charity, government and commercial organizations with an approved UKB project (https://www.ukbiobank.ac.uk/enable-your-research/apply-for-access). The GenoPred Pipeline code is available from the GenoPred GitHub Code Repository (https://opain.github.io/GenoPred/) and our analysis pipeline will be made available on our GitHub page upon publication. GLAD data are available through a data access application to the NIHR BioResource.

## Acknowledgements

We gratefully acknowledge the research participants across all cohorts who generously contributed their time and samples, making this work possible, and the research participants and employees of 23andMe Research Institute for making this work possible. We thank the Estonian Biobank participants and the Estonian Biobank Research Team for providing data. Data analysis was carried out in part at the High-Performance Computing Center of the University of Tartu. UK Biobank analysis was conducted under project 82087.

## AGDS

We thank the participants for their time and support. We also thank those involved in conception, implementation, beta testing, media campaign, and data cleaning. Special thanks go to Katherine Kirk for early project management; Lorelle Nunn, Rebekah Cicero, Mary Ferguson, Lucy Winkler, and Natalie Garden for data and sample collection; Simone Cross for data collection and project management; Lenore Sullivan for participant contact; Natalia Zmicerevska, Alissa Nichles, and Candace Brennan for recruitment support; Jonathan Davies, Luke Lowrey, and Valeriano Antonini for IT support; Vera Morgan and Ken Kirkby for media campaign assistance. Thank you to Scott Gordon for carrying out the imputation and quality control of AGDS data and Penelope Lind for contributing to the management of the AGDS database.

## GLAD and the NIHR Bioresource

We thank the GLAD Study and NIHR BioResource volunteers for their participation, and gratefully acknowledge NIHR BioResource centers, NHS Trusts and staff for their contribution. We thank the National Institute for Health and Care Research, NHS Blood and Transplant, and Health Data Research UK as part of the Digital Innovation Hub Programme. The views expressed are those of the author(s) and not necessarily those of the NHS, the NIHR or the Department of Health and Social Care.

## The AMBER Research Team

Iona Beange^1^, Cristina Douglas^1^, Emer O’Leary^1^, Mark Somerville^1^, Sue Fletcher-Watson^1,2^, Arlene Casey^1^, Matúš Falis^1,3^, Matthew H Iveson^3^, Heather C Whalley^1,3^, Mark J Adams^3^, Megan Calnan^3^, Hannah M Smith^3^, Andrew M McIntosh^3,4^, Solal Chauquet^5^, Zoe L Hunter^5^, Anjali K Henders^5^, Quan Nguyen^5^, Renata Nedel Pertile^5^, Vanessa Tsui^5^, Sonia Shah^5^, Clara Albiñana^6^, Alicia Walker^6^, Naomi Wray^5,6,7^

^1^Usher Institute, University of Edinburgh, Edinburgh, UK

^2^ Salvesen Mindroom Research Centre, University of Edinburgh, Edinburgh, UK

^3^ Institute for Neuroscience and Cardiovascular Research, University of Edinburgh, Edinburgh, UK

^4^Centre for Genomic & Experimental Medicine, Institute for Genetics and Cancer, University of Edinburgh, Edinburgh, UK

^5^ Institute for Molecular Bioscience, University of Queensland, Brisbane, QLD, Australia

^6^ Department of Psychiatry, Oxford University, Oxford, UK

^7^ Queensland Brain Institute, University of Queensland, Brisbane, QLD, Australia

## The AGDS Research Team

Enda M Byrne¹, Jacob J Crouse², Ian B Hickie², Penelope A Lind³,⁴,⁵, Nicholas G Martin⁶, Sarah E Medland³,⁷,⁸, Brittany L Mitchell⁴,⁵,⁹, Richard Parker⁹, Naomi R Wray¹⁰,¹¹

¹ Child Health Research Centre, The University of Queensland, Brisbane, Australia ² Brain and Mind Centre, The University of Sydney, Sydney, Australia

³ Psychiatric Genetics, QIMR Berghofer Medical Research Institute, Brisbane, Australia

⁴ School of Biomedical Sciences, The University of Queensland, Brisbane, Australia

⁵ School of Biomedical Sciences, Queensland University of Technology, Brisbane, Australia

⁶ Genetic Epidemiology, QIMR Berghofer Medical Research Institute, Brisbane, Australia

⁷ School of Psychology, The University of Queensland, Brisbane, Australia

⁸ School of Psychology and Counselling, Queensland University of Technology, Brisbane, Australia

⁹ Brain and Mental Health Program, QIMR Berghofer Medical Research Institute, Brisbane, Australia

¹⁰ Department of Psychiatry, University of Oxford, Oxford, UK

¹¹ Institute for Molecular Bioscience, The University of Queensland, Brisbane, Australia

## GLAD Study Consortium

Gerome Breen¹,²,³, Thalia C Eley¹,³, Chérie Armour⁴, Mark J Adams⁵, Andrew M McIntosh⁵, James TR Walters⁶, Evangelos Vassos¹,³, Matthew Hotopf³,⁷, Esteban Tato-Barcia¹,³, Jonathan RI Coleman¹,²,³

¹ Social, Genetic & Developmental Psychiatry Centre, Institute of Psychiatry, Psychology and Neuroscience, King’s College London, London, UK

² NIHR BioResource, Cambridge University Hospitals, Cambridge Biomedical Campus, Cambridge, UK ³ National Institute for Health and Care Research Biomedical Research Centre: Maudsley, South London and Maudsley NHS Foundation Trust, London, UK

⁴ School of Psychology, Queen’s University Belfast, Belfast, UK

⁵ Institute for Neuroscience and Cardiovascular Research, University of Edinburgh, Edinburgh, UK

⁶ MRC Centre for Neuropsychiatric Genetics and Genomics, Cardiff University, Cardiff, UK

⁷ Department of Psychological Medicine, Institute of Psychiatry, Psychology and Neuroscience, King’s College London, London, UK

## Ethics

All participants provided informed consent, and each cohort received ethical approval from the relevant committee. UK Biobank has approval from the North West Multi-center Research Ethics Committee as a Research Tissue Bank (11/NW/0382, renewed as 16/NW/0274 and 21/NW/0157), and this research was conducted under UK Biobank application 82087. The GLAD Study was approved by the London–Fulham Research Ethics Committee (18/LO/1218), and participants were recruited through the NIHR BioResource, which has approval from the East of England–Cambridge Central Research Ethics Committee (17/EE/0025). The All of Us Research Program is overseen by the All of Us Institutional Review Board, which has determined that research using Controlled Tier data does not constitute human subjects research and does not require individual project review. For cohorts contributing GWAS summary statistics, 23andMe participants in both the Antidepressant Efficacy and Side Effects survey (Li et al., 2016) and the Antidepressant Efficacy survey (Li et al., 2020) provided informed consent and participated in the research online under a protocol approved by an external institutional review board, Ethical & Independent Review Services. The Australian Genetics of Depression Study was approved by the QIMR Berghofer Medical Research Institute Human Research Ethics Committee (P2118). The activities of the Estonian Biobank are regulated by the Human Genes Research Act, and all EstBB participants provided broad informed consent.

## Declaration of generative AI and AI-assisted technologies

During the preparation of this manuscript, the author(s) used AI tools - Claude (Anthropic), to assist with refining, editing and formatting human-written text. The author(s) reviewed and edited all AI-assisted content and take responsibility for the accuracy and integrity of the final manuscript.

